# Implicit, Intrinsic, Extrinsic, and Host Factors Attributing the Covid-19 Pandemic. Part 1- Intrinsic Factor Nitrous Oxide Emissions: A Systematic Analysis

**DOI:** 10.1101/2021.09.07.21263196

**Authors:** Nikita Thapliyal, Gaurav Joshi, Prashant Gahtori

## Abstract

Despite Nitrous oxide (N_2_O) being the most widely used anesthetics in dental and other medical applications, it is associated with global warming and stratospheric ozone destruction. With globalization, a larger amount of N_2_O emissions arearticulated especially from human activities (30%, 6.7 Tg N per year), which are primarily dominated by agriculture that is even above the emissions of all oceans (26%). The synthesis of N_2_O reflects the general chemistry and readily from a substrate Nitric oxide (NO) in the environment. The modeling of infectious disease dynamics covering common pathogen-transmission factors, for example intrinsic (or microbes nutrient supply) at a population level, is indeed imperative to curb the menace of any disease. Nonetheless, in areas where novel coronavirus disease (COVID-19) was at its worst, for example, Wuhan China, Mumbai India, Milan Italy, Washington USA etc., the reduction in N_2_O emissions was well noticed. Nonetheless, viruses exhibit greater mobility than humans and hijack nutrients including nitrogen to complete their epidemiological cycle all due to limited sequence space of viral genomes, the high probability of genetic drift, extremely large population sizes, the high mutation and recombination rates. In consequence of drastic fall in N_2_O emissions, lower human transport can not be an all alone contributor, but contrarily it may also be associated with coronavirus intrinsic factors. This prompted us to analyze freely accessible and large global data from two authenticated sources, the World Health Organization and World Bank. We hereby argue that intrinsic factor N_2_O emissions fueling the COVID-19 progression significantly. Entire predictions were found consistent with the recently observed shreds of evidence. These insights enhanced scientific ability to interrogate viral epidemiology and recommended a 7-points framework covering all-natural lifestyle and dietary supplements for COVID-19 prevention before the arrival of a front-line therapeutic(s) or preventable vaccine.

## Introduction

Over the years, the oxides of nitrogen have become increasingly important in contemporary science and are subjected to many reviews [1,2]. Among them three oxides, Nitrous oxide (N_2_O) popularly known as laughing gas, Nitrogen dioxide (NO_2_) and a free radical molecule Nitric oxide (NO) are the most abundant in the air [3]. N_2_O oxidation by ozone can occur at any temperature and considered as an ozone depleting substance with a long half-life (100 to 150 years). The NO (k ∼10^7^ lit mol^-1^ sec^-1^) is very rapidly oxidized to NO_2_ by ozone. The NO_2_ (k ∼3×10^4^ lit mol^-1^ sec^-1^) is still rapidly but slowly than NO oxidized to NO_3_ by ozone.

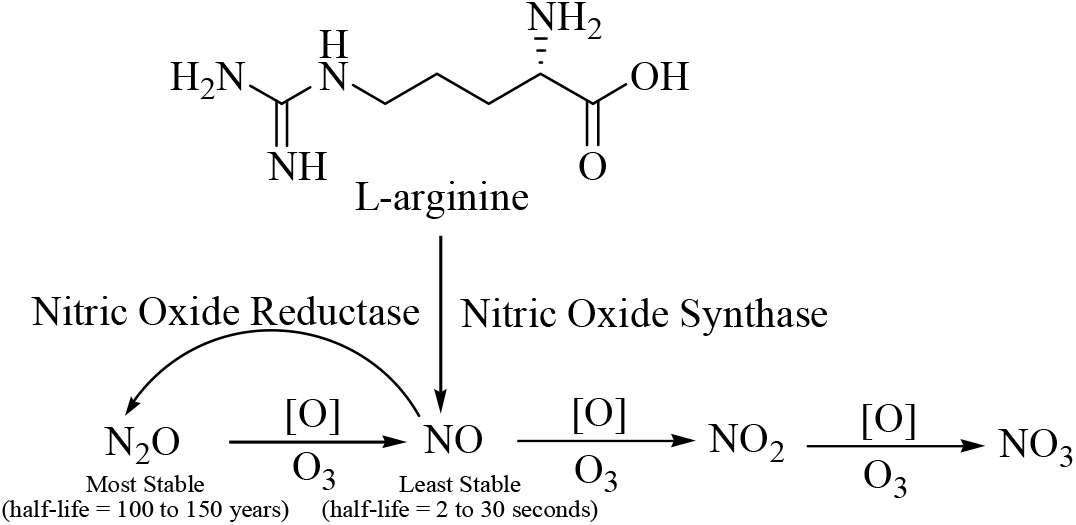

The NO is produced in many tissues from L-arginine and molecular oxygen by four distinct isoforms of Nitric Oxide Synthase (NOS): (i) neuronal NOS-1 (nNOS), (ii) inducible NOS-2 (iNOS), (iii) endothelial NOS-3 (eNOS) and (iv) mitochondrial NOS (mtNOS). Nitric oxide reductases (NOR) play a predominant role in the detoxification of NO and likely to increase fitness of many Gram-positive and – negative pathogens during their associations with invertebrate and vertebrate hosts. The presence of an unpaired electron with uncharged molecule turns NO as an ideal messenger in human with a very short half-life of 2 to 30 seconds [4]. These characteristics of NO also allow its free diffusion across biomembranes and spontaneous signal transmission to perform a wide variety of physiological functions including an olfactory signal procession; platelet clotting; smooth muscle relaxation and vasodilation; blood flow regulation in the brain, heart, lung, gastrointestinal tract, and kidney; blood pressure regulation; cardiac contractility; and killing or inhibition of pathogens etc.[5,6] Despite having large physiological functions, the Global warming potential of 1 gram of N_2_O is 265-298 times of same carbon dioxide for a 100-year timescale. A recent report published in Nature by Tian and coworkers highlighted that N_2_O emissions from human activities such as agriculture, fuel combustion, wastewater management, and industrial processes have ballooned 30% over the past forty years [7]. Here agriculture constitutes the largest source of N_2_O gas emissions, 100-, 8-, and 5-folds in comparison to transport, industry, and international bunkers respectively. Needless to say, that a well-balanced ecosystem is a cornerstone of human health and well-being[8,9]. However, research into how natural environments relates to human health remains limited in important respects.

The COVID-19 disease surge caused by the novel corona (SARS-COV2) virus is posing enormous health challenges since its first detection in China during December 2019, and subsequently its declaration as a pandemic by the World Health Organization (WHO) on dated March 11, 2020 [10]. Recent MEDLINE records of COVID-19 accessed via PubMed shares 0.155931 million results during the writing of the manuscript as on July 13, 2021, which further emphasize several successful scientific accomplishments across its virology, biology, prevention, therapeutics, diagnostics, etc. [11] Despite these success stories, still the eradication of COVID-19 is limited to 3Ts plus P i.e. Tracing (immediate patient isolation), Testing (early diagnosis), Treatment (supportive respiratory care) and Prevention (vaccination). However, according to a recent estimate, there has been a total of 185.29153 million confirmed COVID-19 cases, resulting in 4.010834 million deaths and 0.458355 million new cases are still appearing worldwide on daily basis. [12] The COVID-19 patients have mild to severe symptoms, most of are seeming closely linked with the physiological functions of NO such as loss of olfactory sensation; platelet clotting; difficulty in breathing; multi-organ brain, heart, lung, gastrointestinal tract, and kidney failure etc. [13] In fact, recently the FDA approved an inhaled nitric oxide (iNO plus) for COVID-19 emergency use to a biotherapeutics company, Bellerophon Therapeutics. [14] In addition, a fall in N_2_O gas emissions was noticed in local affected areas after the surge of the SARS-CoV-2 virus. Consequently, these real evidences convince us about the physiological role of substrate NO in COVID-19 management and huge spread of SARS-CoV-2 virus may be due to atmospheric imbalance of N_2_O. Furthermore, there is considerable evidence that intrinsic, extrinsic (or environment), implicit, and host factors are key regulators of any pathogen transmission [15] and an intrinsic factor both operationally and conceptually have yet to be reached for full annihilation of COVID-19. Supressing N_2_O emissions seems the most viable pathway for attaining the great health challenges posed by COVID-19 pandemic. This prompted us to perform a wider sampling of global COVID-19 infectious cases with both nitric oxide and carbon dioxide emissions in understanding the SARS-COV2 transmission in atmosphere and human population.

## Methods

A large authenticated data set with both Covid-19 cases and nitrous oxide/ carbon dioxide emissions consisting 97 countries/ territories is freely accessible, and retrieved from two reliable sources, the World Health Organization (WHO) and the World Bank (n= 97). All COVID-19 infectious cases (n= 97) were collected from five WHO situation reports-dated 09-Jul-2021, 22-Dec-2020, 13-Mar-2020, 19-Mar-2020, and 28-Mar-2020 [16]. All recent value pertaining to both nitrous oxide and carbon dioxide emissions countries/ territories-wise is also publically accessible and supplied by the World Bank [17]. Carbon dioxide emissions listed in per capita were multiplied with country’s population and finaly converted into thousand metric tons. Here, 97 Countries/ Territories randomly fitted to top variable indices of correlation coefficient for unbiased cause-effect relationships (Table 1).

**Table 1.** Cumulative COVID-19 Infected Cases and Nitric oxide/ Carbon dioxideemissionsWorldwide

|  | Country/<br>Territory/Area | CUMULATIVE COVID-19 INFECTED CASES |  |  |  |  | Nitrous Oxide<br>Emissions<br>(Thousand metric<br>tons of CO<sub>2</sub><br>equivalent) | Carbon Dioxide<br>Emissions<br>(Thousand metric<br>tons) |
| --- | --- | --- | --- | --- | --- | --- | --- | --- |
|  |  | 09-Jul-2021 | 22-Dec-2020 | 13-Mar-2020 | 19-Mar-2020 | 28-Mar-2020 |  |  |
| 1 | Albania | 132565 | 52542 | 23 | 59 | 197 | 1,100 | 5,582 |
| 2 | Algeria | 143652 | 94781 | 25 | 72 | 367 | 12,110 | 157,498 |
| 3 | Argentina | 4593763 | 1531374 | 31 | 79 | 589 | 47,930 | 180,206 |
| 4 | Armenia | 226135 | 153825 | 1 | 84 | 372 | 1,170 | 5,572 |
| 5 | Australia | 30905 | 28128 | 140 | 510 | 3635 | 76,760 | 394,624 |
| 6 | Austria | 647111 | 334629 | 361 | 1646 | 7697 | 3,620 | 64,365 |
| 7 | Azerbaijan | 336788 | 199127 | 11 | 34 | 147 | 4,160 | 32,662 |
| 8 | Bahrain | 266797 | 90062 | 195 | 256 | 473 | 90 | 33,339 |
| 9 | Bangladesh | 1000543 | 499560 | 3 | 10 | 48 | 29,240 | 84,459 |
| 10 | Belarus | 424554 | 171579 | 12 | 46 | 94 | 12,010 | 59,096 |
| 11 | Belgium | 1092477 | 625928 | 314 | 1486 | 7284 | 4,460 | 94,800 |
| 12 | Bhutan | 2258 | 446 | 1 | 1 | 3 | 190 | 1,411 |
| 13 | Bolivia | 449687 | 149149 | 3 | 12 | 61 | 8,770 | 23,350 |
| 14 | Brazil | 18909037 | 7162978 | 77 | 291 | 2915 | 179,200 | 434,020 |
| 15 | Brunei<br>Darussalam | 266 | 152 | 12 | 56 | 115 | 150 | 7,282 |
| 16 | Bulgaria | 422353 | 191029 | 7 | 92 | 293 | 4,050 | 40,682 |
| 17 | Burkina Faso | 13505 | 4954 | 2 | 26 | 146 | 9,850 | 4,519 |
| 18 | Cameroon | 80858 | 25849 | 2 | 10 | 75 | 62,710 | 9,075 |
| 19 | Canada | 1418632 | 495346 | 138 | 569 | 4018 | 43,120 | 584,891 |
| 20 | Chile | 1579591 | 583354 | 33 | 238 | 1610 | 6,410 | 88,410 |
| 21 | China | 119141 | 95716 | 80991 | 81174 | 82230 | 538,790 | 10,658,497 |
| 22 | Colombia | 4426811 | 1482072 | 9 | 93 | 491 | 21,720 | 81,446 |
| 23 | Costa Rica | 377091 | 157472 | 22 | 50 | 231 | 1,940 | 8,416 |
| 24 | Cote d Ivoire | 48693 | 21772 | 1 | 9 | 92 | 3,030 | 10,555 |
| 25 | Croatia | 360680 | 194962 | 16 | 81 | 586 | 1,790 | 16,651 |
| 26 | Cyprus | 81581 | 17476 | 6 | 58 | 162 | 370 | 7,340 |
| 27 | Czech Republic | 1668891 | 624140 | 116 | 522 | 2279 | 5,170 | 103,242 |
| 28 | Denmark | 297543 | 131606 | 674 | 1044 | 2046 | 4,740 | 33,372 |
| 29 | Dominican<br>Republic | 331826 | 159064 | 5 | 21 | 581 | 3,390 | 25,642 |
| 30 | Ecuador | 465878 | 205920 | 17 | 155 | 1595 | 4,950 | 40,823 |
| 31 | Egypt | 282582 | 124891 | 67 | 196 | 536 | 22,320 | 256,045 |
| 32 | Estonia | 131396 | 21794 | 13 | 258 | 575 | 1,350 | 16,055 |
| 33 | Finland | 97651 | 32582 | 109 | 359 | 1025 | 4,670 | 44,563 |
| 34 | France | 5686066 | 2418439 | 2860 | 9043 | 32542 | 38,030 | 301,514 |
| 35 | French Polynesia | 19026 | 16182 | 1 | 3 | 30 | 38 | 781 |
| 36 | Georgia | 374836 | 208638 | 25 | 38 | 85 | 2,090 | 9,468 |
| 37 | Germany | 3734468 | 1494009 | 2369 | 8198 | 48582 | 32,900 | 717,056 |
| 38 | Greece | 433021 | 130485 | 98 | 418 | 966 | 4,450 | 63,405 |
| 39 | Guyana | 20645 | 6076 | 1 | 4 | 8 | 970 | 2,464 |
| 40 | Honduras | 270689 | 116212 | 2 | 9 | 67 | 3,210 | 10,093 |
| 41 | Hungary | 808437 | 302989 | 16 | 58 | 343 | 5,790 | 45,843 |
| 42 | Iceland | 6675 | 5621 | 61 | 250 | 890 | 400 | 2,128 |
| 43 | India | 30752950 | 10031223 | 74 | 151 | 724 | 253,790 | 2,459,313 |
| 44 | Indonesia | 2455912 | 657948 | 34 | 227 | 1046 | 95,690 | 589,547 |
| 45 | Iran | 3327526 | 1152072 | 10075 | 17361 | 32332 | 37,580 | 646,159 |
| 46 | Iraq | 1406289 | 583118 | 70 | 164 | 458 | 5,420 | 196,897 |
| 47 | Ireland | 276104 | 78776 | 70 | 292 | 2121 | 10,060 | 37,647 |
| 48 | Israel | 845123 | 371373 | 75 | 427 | 3460 | 2,360 | 60,385 |
| 49 | Italy | 4267105 | 1938083 | 15113 | 35713 | 86498 | 17,900 | 325,065 |
| 50 | Jamaica | 50497 | 12135 | 1 | 13 | 26 | 410 | 8,588 |
| 51 | Japan | 813976 | 195880 | 675 | 873 | 1499 | 18,010 | 1,109,045 |
| 52 | Jordan | 754927 | 272797 | 1 | 52 | 235 | 1,250 | 25,289 |
| 53 | Kuwait | 369227 | 147775 | 80 | 142 | 235 | 750 | 92,341 |
| 54 | Latvia | 137797 | 30297 | 16 | 71 | 280 | 1,860 | 7,468 |
| 55 | Lebanon | 546366 | 156570 | 66 | 133 | 391 | 740 | 27,573 |
| 56 | Lithuania | 279024 | 112359 | 3 | 26 | 358 | 3,530 | 11,262 |
| 57 | Luxembourg | 71879 | 44067 | 17 | 210 | 1605 | 300 | 9,596 |
| 58 | Malaysia | 808658 | 91969 | 129 | 673 | 2161 | 11,120 | 245,989 |
| 59 | Maldives | 74848 | 13474 | 8 | 13 | 13 | 30 | 1,910 |
| 60 | Malta | 30851 | 11621 | 9 | 48 | 139 | 40 | 1,412 |
| 61 | Mexico | 2558369 | 1301546 | 12 | 93 | 589 | 42,250 | 482,399 |
| 62 | Mongolia | 131962 | 953 | 1 | 5 | 12 | 13,190 | 21,320 |
| 63 | Morocco | 537253 | 415226 | 6 | 49 | 358 | 9,430 | 68,311 |
| 64 | Nepal | 652859 | 253184 | 1 | 1 | 3 | 8,330 | 12,476 |
| 65 | Netherlands | 1701911 | 676589 | 614 | 2051 | 8603 | 7,790 | 150,321 |
| 66 | New Zealand | 2409 | 1760 | 5 | 20 | 416 | 15,040 | 31,695 |
| 67 | North Macedonia | 155760 | 77949 | 7 | 36 | 219 | 550 | 7,375 |
| 68 | Norway | 132572 | 42775 | 489 | 1423 | 3581 | 3,190 | 38,119 |
| 69 | Oman | 281688 | 127019 | 18 | 33 | 152 | 780 | 77,580 |
| 70 | Pakistan | 967633 | 454673 | 20 | 241 | 1235 | 60,950 | 212,628 |
| 71 | Panama | 411226 | 206310 | 14 | 86 | 674 | 1,310 | 10,475 |
| 72 | Paraguay | 432801 | 98296 | 5 | 11 | 52 | 10,070 | 8,634 |
| 73 | Peru | 2071637 | 993760 | 22 | 145 | 580 | 9,360 | 55,947 |
| 74 | Philippines | 1455585 | 458044 | 52 | 187 | 803 | 13,640 | 142,241 |
| 75 | Poland | 2880670 | 1202700 | 49 | 287 | 1389 | 21,480 | 311,685 |
| 76 | Portugal | 899295 | 370787 | 41 | 642 | 4268 | 3,090 | 49,358 |
| 77 | Qatar | 222918 | 141858 | 262 | 442 | 562 | 880 | 94,296 |
| 78 | Republic of Korea | 165344 | 49665 | 7979 | 8413 | 9478 | 10,770 | 626,745 |
| 79 | Republic of Moldova | 257272 | 134578 | 4 | 36 | 199 | 1,160 | 12,781 |
| 80 | Russian Federation | 5733218 | 2848377 | 34 | 147 | 1264 | 58,610 | 1,623,065 |
| 81 | Saudi Arabia | 497773 | 360848 | 21 | 238 | 1104 | 5,750 | 531,565 |
| 82 | Senegal | 44790 | 17670 | 10 | 36 | 119 | 7,820 | 10,413 |
| 83 | Slovakia | 391813 | 151336 | 21 | 105 | 295 | 1,970 | 33,078 |
| 84 | Slovenia | 257619 | 105013 | 57 | 286 | 632 | 790 | 14,084 |
| 85 | South Africa | 2135246 | 912477 | 17 | 116 | 1170 | 18,820 | 444,616 |
| 86 | Spain | 3915313 | 1797236 | 2965 | 13716 | 64059 | 20,350 | 258,103 |
| 87 | Sri Lanka | 269946 | 36049 | 3 | 42 | 106 | 2,240 | 21,374 |
| 88 | Sweden | 1092308 | 367120 | 620 | 1279 | 3046 | 4,920 | 35,731 |
| 89 | Switzerland | 701473 | 402264 | 858 | 3010 | 12104 | 2,140 | 38,098 |
| 90 | Thailand | 317506 | 4331 | 75 | 212 | 1136 | 19,950 | 259,240 |
| 91 | Togo | 14176 | 3396 | 1 | 1 | 25 | 1,870 | 2,372 |
| 92 | Tunisia | 464914 | 119151 | 7 | 29 | 227 | 3,320 | 30,637 |
| 93 | Turkey | 5465094 | 1189947 | 1 | 191 | 5698 | 34,380 | 422,996 |
| 94 | Ukraine | 2240246 | 964448 | 3 | 16 | 311 | 25,310 | 181,678 |
| 95 | United Kingdom | 5022897 | 2004223 | 594 | 2630 | 14547 | 28,290 | 1,411,859 |
| 96 | United States of America | 33451965 | 17314834 | 1264 | 7087 | 85228 | 250,060 | 1,786,987 |
| 97 | Vietnam | 24810 | 1411 | 39 | 66 | 169 | 24,080 | 262,698 |
| Correlation | Nitrous Oxide Emissions | 0.6 | 0.5 | 0.7 | 0.7 | 0.6 |  |  |
|  | Carbon Dioxide Emissions | 0.2 | 0.2 | 0.9 | 0.9 | 0.6 |  |  |

\*Top with red, Bottom with green

## Results and Discussion

We ascertained a large data set involving a total of ninty seven Countries/ Territories randomly who fulfilled the unbiased study criteria used in the assessment of role of nitrous oxide emissions over Covid-19 pandemic. There was a large variation among both N_2_O emissions (variation= 4.53 × 10^9^) and cumulative COVID-19 cases (variation ranges from 2.48×10^13^ to 2.88×10^8^ dated from 28-March-2020 to 09-July-2021) across all Countries/ Territories (Table 1). Here with the large variation, we observed a good and consistent correlation (r^2^= 0.6 ± 0.06) between N_2_O emissions and Cumulative covid-19 cases from last one year plus (Table 2). Moreover, the Z-test with a probability, P values falls below the level of significance at 0.05 (5%) (one-tail ranges from 0.0002 to 0.0034 and two-tail ranges from 0.0004 to 0.0068). This reveals that there is a significant relationship between N_2_O emissions and Cumulative COVID-19 cases. The result of the analysis shows the calculated values of the Z-test of 3.2776 (22-Dec-2020)and 3.5593 (09-July-2021) which are higher than the Z-critical two-tail value of 1.9600 revealing that there is a positive significant relationship between N_2_O emissions and cumulative covid-19 cases. Other hands, in the case of CO_2_ emissions, a weak and inconsistent correlation (r^2^= 0.2 ± 0.23) exist, suggesting that COVID-19 cases are independent of CO_2_ utilization. These statistical findings were overall supportive of N_2_O emission’s impact on enhancing SARS-COV2 spread in the human population.

**Table 2.** z-Test: Two sample for means (Cumulative COVID-19 infected cases and Nitrous oxide emissions)

|  | CUMULATIVE COVID-19 INFECTED CASES |  |  | Nitrous Oxide Emissions |
| --- | --- | --- | --- | --- |
|  | 09-Jul-2021 | 22-Dec-2020 | 28-Mar-2020 |  |
| Mean | 1825200.4 | 740951.0309 | 5815.2062 | 24886.4 |
| Known Variance | $2.48 \times 10^{13}$ | $4.63 \times 10^{12}$ | $2.88 \times 10^8$ | $4.53 \times 10^9$ |
| Observations | 97 | 97 | 97 | 97 |
| Hypothesized Mean Difference | 0 | 0 | 0 |  |
| z | 3.5593 | 3.2776 | -2.7060 |  |
| P(Z<=z) one-tail | 0.0002 | 0.0005 | 0.0034 |  |
| z Critical one-tail | 1.6449 | 1.6449 | 1.6449 |  |
| P(Z<=z) two-tail | 0.0004 | 0.0010 | 0.0068 |  |
| z Critical two-tail | 1.9600 | 1.9600 | 1.9600 |  |

Notably, the top five N_2_O emissions countries China, India, United States of America (US), Brazil, and Indonesia. [18] According to the WHO, currently the US, India and Brazil are the top three countries with the highest number of infections in the world. Yet standing at the 15th global rank of COVID-19, in our opinion Indonesia is still at high risk. Other hand, the Brunei Darussalam, Maldives, French Polynesia, Malta, Bahrain and Bhutan are among the least N_2_O emissions Countries/ Territories. All these are with the least COVID-19 outbreaks. Overall, these findings are further supported by real global evidences and suggest N_2_O gas emissions significantly increase SARS-COV2 virus transmission.

Statistical analyses of epidemiological data summarize the way by which COVID-19 spread in the human population. According to the Oxford University report, agriculture constitutes the largest source of N_2_O gas emissions 100-folds in comparison to transport vehicles, and both synthetic fertilizers/pesticides usually increase the negative environmental footprint of agriculture by contributing to greenhouse N_2_O emissions. Surprisingly, a fall in N_2_O gas emissions was noticed in affected areas after the surge of the SARS-CoV-2 virus [19] and is supportive of a natural virus evolution utilizes the major N_2_O emissions.

By science value, there is a growing awareness that any infectious disease modulate the interplay between pathogens, hosts and environment. As stated by the National Institutes of Health that once any pathogen withstand the environment outside its host for a long period of time, an indirect contact occurs and this type of indirect transmission is already well established in case of gastrointestinal diseases, for example cholera, rotavirus infection, cryptosporidiosis, and giardiasis [20]. Hence, these sporadic COVID-19 instances are in full agreement with intrinsic N_2_O emissions and suggest that these emissions is emerging as SARS-COV2 virus feed supplements.

Nature remains silent killer of harmful pathogens, and at the core of its response is the fundamental realization that nature’ effect against COVID-19 epidemic is varied around the globe. Consequently, policy and strategy response covering supression of N_2_O emissions underlying scientific interventions for effective tackling of COVID-19 pandemic before arrival of front-line therapeutics and preventive vaccine.

The COVID-19 attributing factors with recently observed evidence, for instance, (i) a fall in N_2_O gas emissions after the surge of COVID-19 in China, N_2_O itself is a biochemical precursor of NO (↓NO level); (ii) High blood sugar-a reduced ability to produce NO (↓NO level); (iii) High blood pressure-NO inactivation (↓NO level); (iv) Obesity inhibit NO (↓NO level); (v) Smoking-inhibit biomolecular signaling molecule NO via nicotinic acetylcholine receptors (↓NO level); (vi) concurrent drug use (↓NO level); and (viii) a high salt intake attenuates NO production (↓NO level). It has been corroborated that the SARS-COV2 virus is surviving using N_2_O energy source in atmosphere and in human population. Once again, COVID-19 pathophysiology and common symptoms fever, apnoea, cough, tiredness, and highly vulnerable groups, such as hypertensive, diabetics, smoker, and obese people, all observed reduced bioactive NO after the COVID-19 viral entry. [21]

NO is a gaseous secondary messenger known to be involved in numerous intra- and intercellular signaling events[22-24], for example, regulation of non-specific immune defense, body temperature, blood sugar, inflammation, vasodilation, airway tone, etc. More specifically, reports by Dantonio et al. suggested systemic inhibition of NO synthesis attenuates bacterial endotoxin-induced fever[25], Hori et al. highlighted NO is involved in an ovalbumin-induced increase in cough reflex sensitivity[26], Prado et al. outlined role of nitric oxide in the various pathologic mechanisms of allergic asthma[27]. As a result of NO inhibition, common symptoms fever, cough, tiredness, and difficulty in breathing were appearing after COVID infection. The NO inducible agents, for example, hydroxychloroquine is being clinically relevant in COVID-19 treatment. A review of Nitric oxide enhancement strategies emphasizes many dietary strategies for more safe and effective NO based solutions than NO donating therapeutics[28].

The NO inactivation concept has evolved from the current approach and, endorse novel SARS-CoV2 coronavirus is greatly dependent upon local places with high N_2_O emissions in past and CURRENT N_2_O is turning low. It’s surprising but most infected places, Wuhan, New York, Mumbai, Milan etc. are perfect falling in this line (Intrinsic-factors). In response, this study also proposed a framework of all-natural supplements for COVID-19 reduction (not prevention) by means of lifestyle and dietary supplements as follows:

1. Smoker, Hypertensive, Diabetics, and Obese people are among highly vulnerable groups, require counselling and strict attention (As reported above).
2. Indoor fitness exercises including meditation/ yoga are one of the front-line COVID-19 reduction markers. (Increase NO production).
3. People are advised to consume a very minimum amount of sugar and salt. (Both salt and sugar decreases NO production).
4. Patients with poly-pharmacy or concurrent drugs, for example, statins, fibrates, thiazolidinediones, metformin, antioxidant vitamins, aspirin, n-3 polyunsaturated fatty acids, and plant flavonoids should remain attentive. (Poly-pharmacy attenuate NO production).
5. Consumption of natural L-Arginine-rich sources, for example, dairy products, soybeans, peanuts, spirulina, lentils, eggs, etc. (L-Arginine is a precursor of NO production) and Citrulline supplementation using natural sources, for example, watermelon, pumpkins, cucumber, etc. (Citrulline stimulates de novo Arginine production)
6. Moderate consumption of nitrate-rich natural sources, such as beetroot, spinach, lettuce, chervil, radish, celery, pomegranate, etc. (Nitrate-nitrite-NO is second metabolic pathway).

In line with modern science multi-factorial approach, this study is endorsing nature’s role in direct pathogen invasion and to limit the transmission of SARS-COV-2 pathogen. Thus, similar approach help researchers think undoubtedly about the causation of COVID-19 disease. We hope the insights presented above along with disease reduction framework will be a game-changer in fighting against the COVID-19 pandemic before the launch of a front-line therapeutic(s) or preventable vaccine.

## Data Availability

WHO and Worldbank

https://www.who.int/emergencies/diseases/novel-coronavirus-2019/situation-reports

## Acknowledgements

The authors are thankful to the World Health Organization and Worldbank for keeping data free to use. The authors are also thankful to GEHU India for providing necessary facilities and technical support.

## Author contributions

PG devised the project and the main conceptual ideas. NT drafted the manuscript. GJ critically revised the manuscript and all authors approved the final version for submission.

